# Genomic epidemiology of SARS-CoV-2 reveals multiple lineages and early spread of SARS-CoV-2 infections in Lombardy, Italy

**DOI:** 10.1101/2020.07.19.20152322

**Authors:** Claudia Alteri, Valeria Cento, Antonio Piralla, Valentino Costabile, Monica Tallarita, Luna Colagrossi, Silvia Renica, Federica Giardina, Federica Novazzi, Stefano Gaiarsa, Elisa Matarazzo, Maria Antonello, Chiara Vismara, Roberto Fumagalli, Oscar Massimiliano Epis, Massimo Puoti, Carlo Federico Perno, Fausto Baldanti

## Abstract

From February to April, 2020, Lombardy (Italy) was the area who worldwide registered the highest numbers of SARS-CoV-2 infection. By extensively analyzing 346 whole SARS-CoV-2 genomes, we demonstrated the simultaneous circulation in Lombardy of two major viral lineages, likely derived from multiple introductions, occurring since the second half of January. Seven single nucleotide polymorphisms (five of them non-synonymous) characterized the SARS-CoV-2 sequences, none of them affecting N-glycosylation sites. These two lineages, and the presence of two well defined clusters inside Lineage 1, revealed that a sustained community transmission was ongoing way before the first COVID-19 case found in Lombardy.

## Introduction

Since coronavirus disease 2019 (COVID-19), was initially reported in China on 30th December 20191,2, SARS-CoV-2 spreads world-wide and, as of 14^th^ June 2020, there have been 7.55 million confirmed infections and 423,000 deaths reported worldwide (World Health Organization, 2020). In Italy, the first case of evident SARS-CoV-2 transmission occurred at February 20^th^, in Codogno, Lombardy, when a young man affected by an interstitial pneumonia was diagnosed for SARS-CoV-2. Since that day, the number of diagnosed COVID-19 cases exponentially increased and Lombardy, become the area most affected by the COVID-19 pandemic, with a total number of 89,018 infections at June 2, out of 233,197 total cases in Italy. Thereafter, the COVID-19 epidemic grew exponentially during the first days of March 2020, peaking on 21 March 2020 with 6,557 newly confirmed cases per day. Two months later, reported COVID-19 cases in Italy dropped to ∼600 per day, indicating the epidemic is close to be contained.

Lombardy is the most populated region in Italy (10 million inhabitants) and one of the widest. Milan represents the largest metropolitan area in Italy and the third most populated functional urban area in Europe with a strong economical and transportation links to Europe and outside. This scenario makes Lombardy the perfect place to host and favor the spread of a highly transmissible virus, such as SARS-CoV-2.

In this study, an integrated approach including epidemiological and viral genetic data was used to reconstruct the pattern of SARS-CoV-2 spread in this region. Whole genome sequencing was performed for 346 SARS-CoV-2 strains obtained from individuals of different geographical areas and in a time span of 2 months. The main aim of this study was to trace the transmission chains and the temporal and geographical evolution of the virus in Lombardy also in relation to the measures implemented to contain it.

## Methods

### Sample collection, and epidemiological data

This retrospective cohort study included 371 SARS-CoV-2-positive nasopharyngeal-swabs of adult patients hospitalized or referred for the diagnosis at two major Hospitals in Lombardy since February 22, to April 4. Clinical data were obtained retrospectively by pseudonymised electronic medical records. The study protocol was approved by local Research Ethics Committee of the two hospitals (prot. 92-15032020 and P_20200029440). This study was conducted in accordance with the principles of the 1964 Declaration of Helsinki.

The severity of the disease was classified into mild, moderate, or severe, if showing i) mild clinical symptoms without sign of pneumonia on imaging, ii) fever and respiratory symptoms with radiological findings of pneumonia, iii) respiratory distress, with oxygen saturation ≤93% at rest, mechanical ventilation, or presence of multiorgan failure (septic shock) and/or admission to intensive care unit (ICU) hospitalization.

### Virus amplification and sequencing

Total RNAs were extracted from nasopharyngeal swabs by using QIAamp Viral RNA Mini Kit, followed by purification with Agencourt RNAClean XP beads. Both the concentration and the quality of all isolated RNA samples were measured and checked with the Nanodrop. Virus genomes were generated by using version 1 of the CleanPlex SARS-CoV-2 Research and Surveillance Panel (https://www.paragongenomics.com/product/cleanplex-sars-cov-2-panel/), according to the manufacturer’s protocol starting with 50 ng total RNA and followed by Illumina sequencing on a NextSeq 500. Briefly, the multiplex PCR was performed with 2 pooled primer mixture and the cDNA reverse transcribed with random primers was used as a template. After 10 rounds of amplification, the two PCR products were pooled and purified. Then the digestion reaction was performed to remove non-specific PCR products, followed with second PCR reaction for barcoding with 24 rounds of amplification. Libraries were checked using High Sensitivity Labchip and quantified with Qubit. Equimolar quantity of libraries was pooled, and the obtained run library mix was loaded at 1.5pM into NextSeq500 for sequencing in the Mid Output format with paired-end 2 x 150 bp. The Illumina sequencing platform takes less than 26 hours to obtain 30.2 Gb of sequencing data (compressed format), achieving between 170.000 and 856.000 paired-end fragments per sample (340.000 and 1.712.000 sequences), with a mean coverage depth of 2.500.

### Virus genome assembly

Reference-based assembly of the metagenomic raw data was performed as follows. Illumina adapters were removed, and reads were filtered for quality (average q28 threshold and read length > 135 nt) using FASTP.^3^ First and last 15 nucleotides were then removed from all reads. The mapping of cleaned reads was performed against the GenBank reference genome NC_045512.2 (Wuhan, collection date: December 2019) using BWA-mem^4^, and consensus sequences were generated using samtools 1.10.^5^ Single nucleotide polymorphisms (SNP variants) were called through a pipeline based on samtools/bcftools^6^, and all SNPs having a minimum supporting read frequency of 40% with a depth ≥100 were retained in the consensus sequence.

### Phylogenetic analysis

The consensus sequence obtained merging the most represented SNP (intra-patient prevalence >40%) was retained per patient. Moreover, all available whole-genome SARS-CoV-2 sequences (n = 3244) on GISAID (gisaid.org) on 3 May 2020 were downloaded. Sequences from GISAID that were error-rich, those without a date of sampling and identical sequences from each country outbreak were removed. Finally, the dataset was reduced by only retaining the earliest, and the most recently sampled sequences from each country outbreak (range of dates: December, 24 2019 – April, 4 2020). The resulting dataset of 205 GISAID sequences therefore represents the global diversity of the virus while minimizing the impact of sampling bias. The 205 Gisaid deposited sequences were added to the consensus sequence obtained by our samples. Sequences were aligned using ClustalX and manually inspected in Bioedit. The final alignment length was 29,282 nucleotides. We used both the maximum likelihood (ML) and Bayesian coalescent methods to explore the phylogenetic structure of SARS-CoV-2. The ML phylogeny was estimated with RAxML^7^ using the under the best-fit model of nucleotide substitution GTR+I^8^ with gamma-distributed rate variation^9^. Tree topology was assessed with the fast bootstrapping function with 1000 replicates. The ML tree was inspected in TempEst^10^, in order to define the correlation between genetic diversity (root-to-tip divergence) and time of sample collection (Supplementary Figure 1). In order to obtain a corresponding time-scaled maximum clade credibility tree, a Bayesian coalescent tree analysis was undertaken with BEAST v1.10.4,^11^ using the HKY+Q4 substitution model with gamma-distributed rate variation with an exponential population growth tree prior and an uncorrelated relaxed molecular clock, under a noninformative continuous-time Markov chain (CTMC) reference prior^12^. Taxon sets were defined and used to estimate the posterior probability of monophyly and the posterior distribution of the tMRCA of observed phylogenetic clusters. Four independent chains were run for 50 million states and parameters and trees were sampled every 1,000 states. Upon completion, chains were combined using LogCombiner after removing 10% of states as burn-in and convergence was assessed with Tracer. The maximum clade credibility (MCC) tree was inferred from the Bayesian posterior tree distribution using TreeAnnotator, and visualized with FigTree 1.4.4 (http://tree.bio.ed.ac.uk/software/figtree/). Monophyly and tMRCA (time to the most recent common ancestor) statistics were calculated for each taxon set from the posterior tree distribution.

### Statistical Analysis

Data were analyzed using Rgui and the statistical software package SPSS (v32.0; SPSS Inc., Chicago, IL).

### Data availability

Sequences are in the process to be deposited in Gisaid (https://www.gisaid.org/) and will be completed by acceptance of the manuscript. Additional data that support the findings of this study are available from the authors upon request.

## Results

### Patients’ characteristics

From February 22 through April 4, 2020, nasopharyngeal swabs of a total of 25,082 patients were screened for SARS-CoV-2 infection at two major Hospitals in Lombardy. Of them, 11,445 received a diagnosis of COVID-19. Whole genome sequencing was performed in a total of 371 samples collected from 371 patients residing in all 12 provinces of Lombardy and with varying disease symptoms, ranging from mild to severe. Twenty-five samples were excluded due to failed amplification (n=9), or poor genomic coverage (<60%, n=16). The final study population thus consisted of 346 patients, whose demographic and clinical characteristics are reported in Table 1. This study population was well representative of the whole Lombardy region, with exception of the East part and North valleys (i.e. Brescia, Mantua, Valtellina and Valcamonica, Figure 1). Two-hundred patients (57.8%) were male, and the median age was of 72 (IQR: 53-83) years. Fever was the most common COVID-19 symptom at admission, followed by cough and dyspnea. Chest radiographs or CT scan confirmed a classical bilateral interstitial pneumonia for 24.1% of them. Patients with severe COVID-19 manifestation more frequently suffered from at least one chronic comorbidity, compared to those with moderate and mild manifestations (p=0.002), with greater impact played by hypertension and chronic kidney disease (p<0.001).

**Table 1.** Demographic, and clinical findings of the 346 SARS-CoV-2 infected patients according with lineages observed by ML tree.

|  |  | Overall,<br>N=346 | Lineages<sup>a</sup> |  | P-value<sup>b</sup> |
| --- | --- | --- | --- | --- | --- |
|  |  |  | 1,<br>N=211 | 2,<br>N=131 |  |
| Demographics and clinical characteristics |  |  |  |  |  |
| Age, years |  | 72 (53-83) | 70 (53-83) | 73 (55-83) | 0.737 |
| Sex, Male |  | 196 (57.8) | 109 (51.6) | 85 (64.9) | **0.045** |
| Residency |  |  |  |  |  |
|  | *Milan* | 75 (21.7) | 49 (23.2) | 25 (19.1) | 0.442 |
|  | *Como* | 68 (19.7) | 28 (13.3) | 40 (30.5) | **<0.001** |
|  | *Pavia* | 61 (17.6) | 42 (19.9) | 19 (14.5) | 0.246 |
|  | *Bergamo* | 36 (10.4) | 14 (6.6) | 22 (16.8) | **0.005** |
|  | *Lecco* | 30 (8.7) | 10 (4.7) | 20 (15.3) | **0.002** |
|  | *Lodi* | 32 (9.2) | 30 (14.2) | 1 (0.8) | **<0.001** |
|  | *Cremona* | 26 (7.5) | 23 (10.9) | 2 (1.5) | **0.002** |
|  | *Other<sup>c</sup>* | 18 (5.2) | 15 (7.1) | 2 (1.5) | **0.040** |
| Chronic comorbidities<sup>d</sup> |  | 163 (52.8) | 96 (50.8) | 65 (55.6) | 0.488 |
|  | *Hypertension* | 33 (10.7) | 22 (11.6) | 10 (8.5) | 0.505 |
|  | *Obesity* | 21 (6.8) | 11 (5.8) | 10 (8.5) | 0.363 |
|  | *Diabetes* | 37 (12.1) | 16 (8.5) | 21 (17.9) | **0.022** |
|  | *Cardiovascular disease* | 96 (31.1) | 53 (28.0) | 41 (35.0) | 0.199 |
|  | *Chronic obstructive lung disease* | 43 (13.9) | 22 (11.6) | 21 (17.9) | 0.169 |
|  | *Malignancies* | 39 (12.6) | 24 (12.7) | 14 (12.0) | 0.992 |
|  | *Chronic kidney disease* | 24 (7.8) | 12 (6.3) | 11 (9.4) | 0.447 |
|  | *Chronic liver disease* | 5 (1.6) | 3 (1.6) | 2 (1.7) | 0.935 |
|  | *Other<sup>e</sup>* | 28 (9.1) | 18 (9.5) | 10 (8.5) | 0.933 |
| Symptoms at admission<sup>f</sup> |  |  |  |  |  |
|  | *Fever* | 107 (59.4) | 70 (56.0) | 35 (67.3) | 0.220 |
|  | *Cough* | 64 (27.0) | 42 (32.6) | 21 (40.0) | 0.268 |
|  | *Dyspnea* | 48 (26.7) | 29 (23.2) | 18 (34.6) | 0.168 |
| Time from symptoms-onset to SARS-CoV-2 diagnosis, weeks |  | 0.29 (0.0-0.57) | 0.28 (0.14-0.57) | 0.43 (0.07-0.71) | 0.157 |
| Collection date (month, day) |  | 03-14<br>(03-06; 03-20) | 03-11<br>(03-05; 03-20) | 03-16<br>(03-09; 03-21) | **0.023** |
| Disease severity<sup>g</sup> |  |  |  |  |  |
|  | *Mild* | 168 (70.9) | 112 (74.3) | 54 (65.1) | 0.187 |
|  | *Moderate* | 38 (13.9) | 25 (11.8) | 13 (15.7) | 0.709 |
|  | *Severe* | 31 (11.3) | 14 (6.6) | 16 (19.3) | 0.081 |
| Evidence of Interstitial Pneumonia<sup>h</sup> |  | 56 (24.1) | 30 (20.3) | 25 (30.9) | 0.103 |
| SARS-CoV-2 rtPCR |  |  |  |  |  |
| Mean cycle thresholds<sup>i</sup> |  | 18.7 (16.7-20.0) | 19.0 (17.2-20.2) | 18.1 (16.3-19.9) | 0.060 |
| SNP in SARS-CoV-2 genome |  |  |  |  |  |
| 20268, A to G, syn (nsp15) |  | 12 (3.5) | 12 (5.7) | 0 (0.0) | **0.013** |
| 23575, C to T, non-syn T to I (S) |  | 23 (6.6) | 23 (10.9) | 0 (0.0) | **0.001** |
| 26530, A to G, non-syn D to G (M) |  | 20 (5.8) | 20 (9.5) | 0 (0.0) | **<0.001** |
| 28881-28883, GGG to AAC, non-syn RG to KR (N) |  | 130 (3.8) | 2 (0.9) | 128 (97.7) | **<0.001** |

Data are expressed as median (IQR), or N (%). <sup>a</sup>A total of 343 SARS-CoV-2 isolates are involved in lineages. <sup>b</sup>P-values were calculated by Mann-Whitney test, or Chi<sup>2</sup> test, as appropriate. <sup>c</sup>Other includes Brescia, Mantua, Monza and Brianza, Sondrio and Varese. <sup>d</sup>Data available for 309 patients. <sup>e</sup>Including: Crohn's disease (n=1), Hashimoto's thyroiditis (n=3), familial lipid disorders (n=9), rheumatoid
arthritis (n=3), Amyotrophic Lateral Sclerosis (n=1), cognitive disorders (n=11). <sup>f</sup>Data available for 180 patients. <sup>g</sup>Data available for 237 patients. <sup>h</sup>Diagnosed by X Ray or CT Scan. Data available for 232 patients. <sup>i</sup>Real-time reverse transcription PCR Ct (cycle threshold) values of these samples ranged from 9 to 35 (GeneFinder™ COVID-19 Plus RealAmp Kit, ELITech; Allplex™ 2019-nCoV Assay, Seegene; Corman VM, Landt O, Kaiser M, et al. Detection of 2019 novel coronavirus (2019-nCoV) by real-time RT-PCR. Euro Surveill. 2020;25(3):2000045. doi:10.2807/1560-7917.ES.2020.25.3.2000045; https://www.who.int/docs/default-source/coronaviruse/protocol-v2-1.pdf).

**Figure 1.**
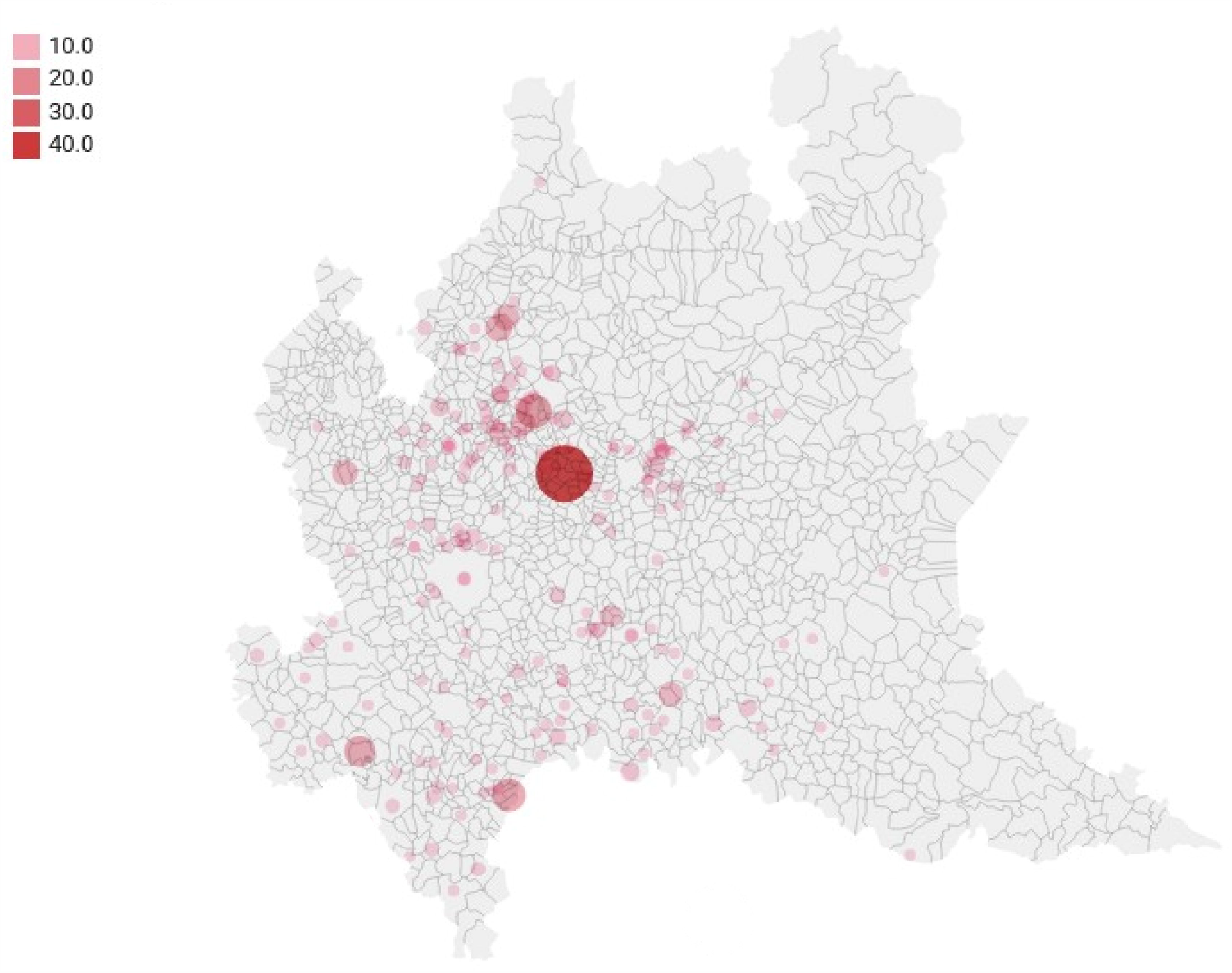
Geographic distribution of COVI D-19 cases among the 12 provinces of Lombardy. Milan: 75; Como: 68; Pavia: 61; Bergamo: 36; Lecco:30; Lodi:32; Cremona: 26; Other (including Brescia, Mantua, Monza and Brianza, Sondrio and Varese): 18

### Genome Coverage

SARS-CoV-2 sequence reads were able to cover from 94.0% to 99.7% of the reference genome (GenBank: NC_045512.2), independently from SARS-CoV-2 load (Supplementary Figure 2). The few genome regions (N=4) with lower reads coverage were consistently limited to no more than 35 nt.

### Single nucleotide polymorphisms characterizing North Italian sequences

The genetic pairwise distance of the 346 sequences indicated that the SARS-CoV-2 sequences evolved progressively during time (rho=0.465, Supplementary Figure 1 and 3). Seven single nucleotide polymorphisms (SNP) were shared among >5 genomes, with a prevalence ranging from 3.6% (20268, A to G, syn in nsp15) to 99.2% (14408, C to T, non-syn P to L in RdRp) (Figure 2 A and B). All SNPs have been detected in at least one previously published SARS-CoV-2 sequence in GISAID. Five out of these seven SNPs were non-synonymous, and three mapped within SARS-CoV-2 structural proteins. Notably, 2 non-synonymous SNPs reside in S protein: the C to T at position 23575 (S protein; amino acid T to I; intra-patient prevalence ∼100%) found in 23 North Italian sequences and previously detected in less than 1% of samples in China, Europe and North America, and the A to G at position 23403 (S protein; amino acid D to G; intra-patient prevalence ∼54%), present in 67.8% of North Italian sequences and firstly detected in China with a prevalence of 1.7%, then rapidly selected and spreading in all countries with a prevalence ranging from ∼17% in Asia to ∼70% in Europe and North America.

**Figure 2.**
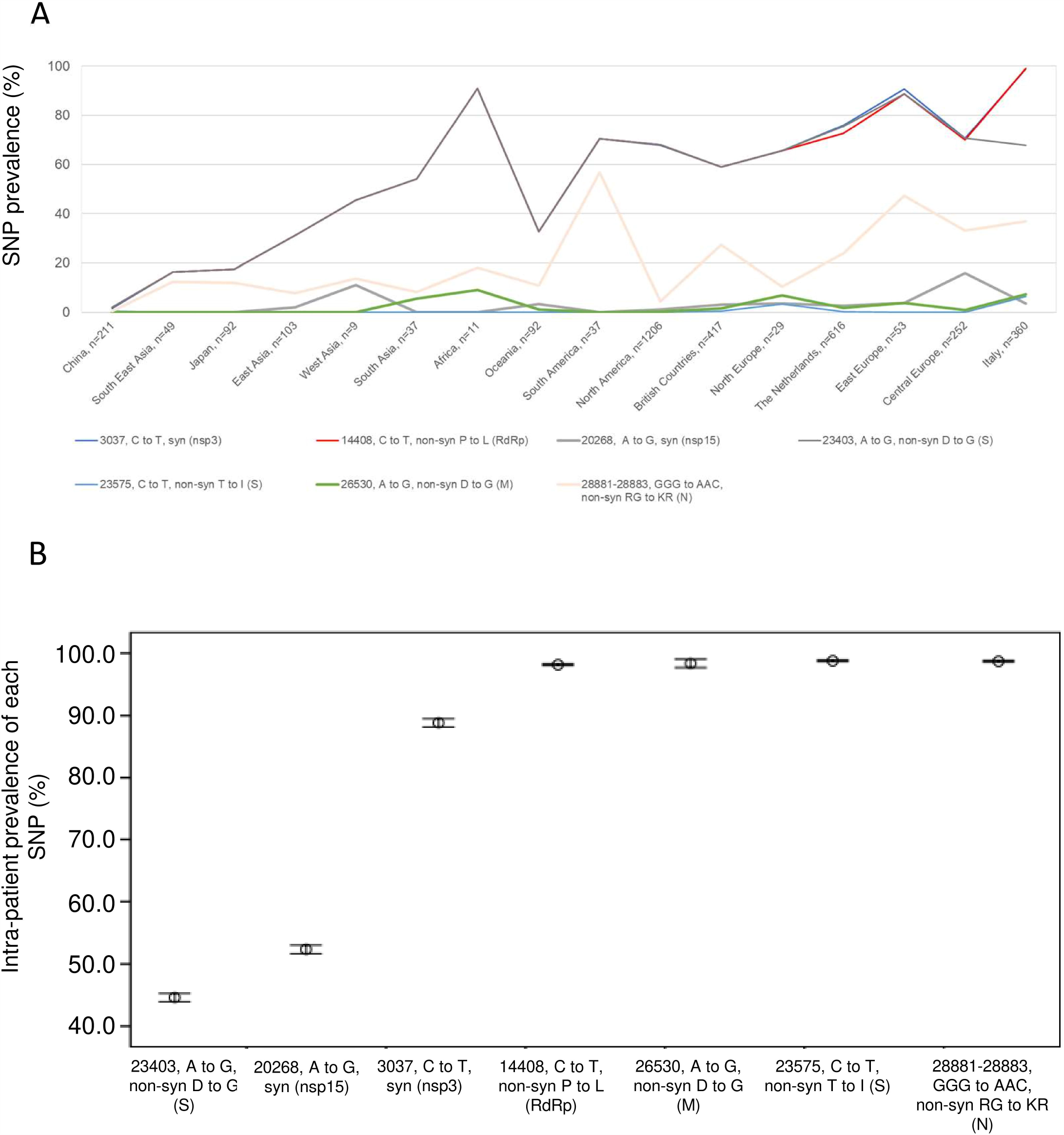
Prevalence of most representative single nucleotide polymorphisms (SNP) in SARS-CoV-2 genomes isolated in Lombardy according to geographical locations and their intra-patient prevalence. (A) Frequency of single nucleotide polymorphisms (with respect to the Wuhan reference genome NC_045512.2) among SARS-CoV-2 sequences according to different geographical location. (B) Intra-patient prevalence of most representative SNPs in North Italian SARS-CoV-2 sequences according to sample date. Italian sequences in panel A include the 6 and 8 sequences from North and Central Italy present in Gisaid at May 05.

### Phylogenetic estimates and lineages characteristics

To explore the distribution of SARS-CoV-2 sequences in Lombardy we performed both an estimated maximum likelihood (ML) phylogeny (Figure 3), and a Bayesian molecular clock analysis (Figure 4). ML tree showed that most of SARS-CoV-2 sequences from Lombardy (grey taxa with red dots, 342/346 [98.8%]) are interspersed within two major viral lineages (1, 2) containing previously isolated Italian strains, and strains from other European, American and Asian countries (gray branches, no dots) (Figure 3). Demographic and clinical characteristics of patients infected with SARS-CoV-2 from such lineages are reported in Table 1.

**Figure 3.**
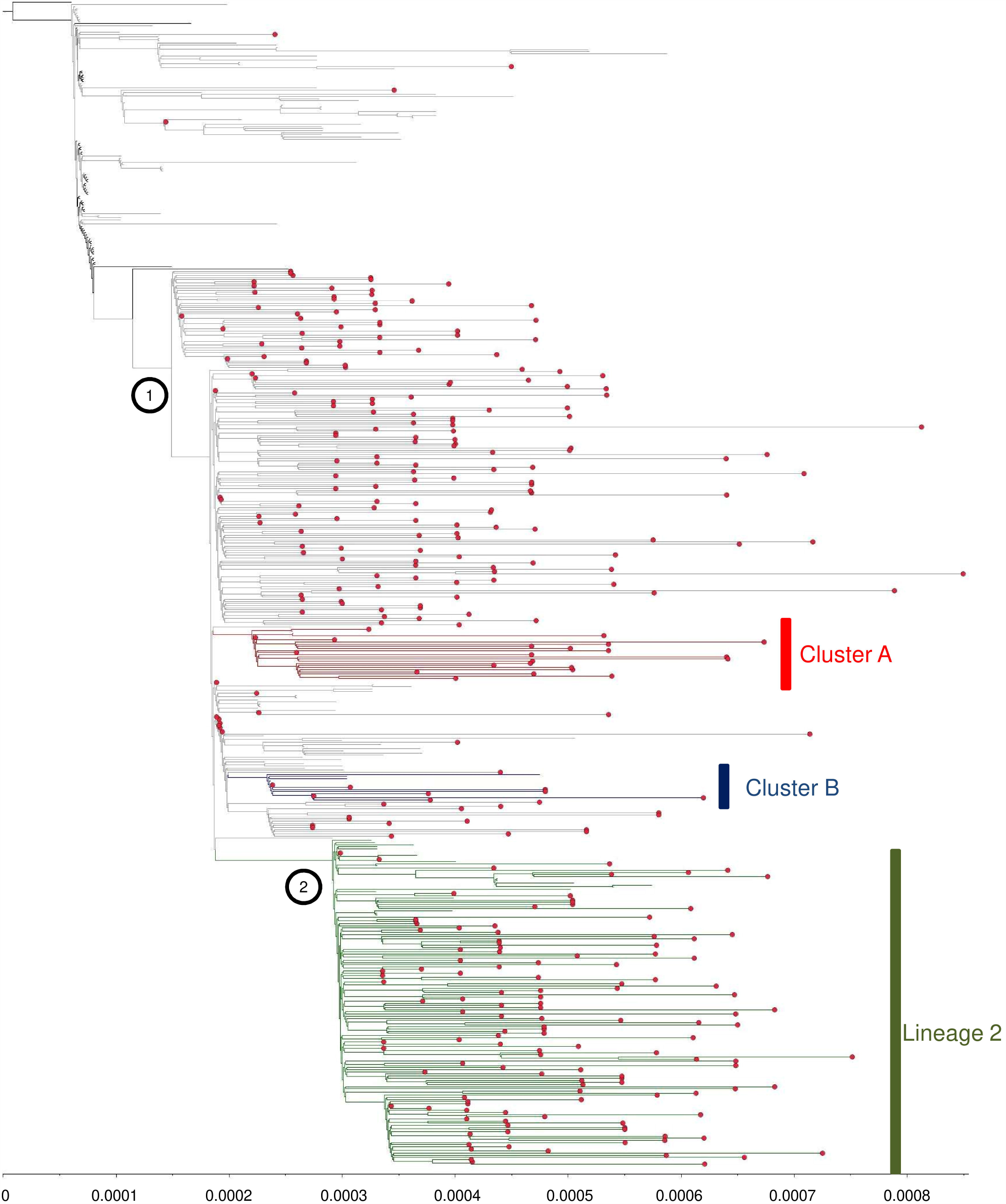
Estimated maximum likelihood phylogeny of SARS-CoV-2 sequences from Lombardy (gray taxa with red dots) and genomes from China (black taxa without dots) and other countries (gray taxa without dots). The two lineages (1 and 2) were reported by black dots at corresponding nodes. SARS-CoV-2 clades supported by a posterior probability of 1.00 in the maximum clade credibility tree were highlighted in green (Lineage 2), in red (cluster A), in blue (Cluster B). Gisaid sequences come from Central Italy (n=6), East Europe (n=6), North Europe (n=6), South America (n=6), Africa (n=7) Japan (n=7), Oceania (n=7), West Asia (n=7), North Italy (n=9), South Asia (n=9), Central Europe (n=11), East Asia (n=11), European Low Countries (n=12), South East Asia (n=13), North America (n=18), British Countries (n=26), China (n=36).

**Figure 4.**
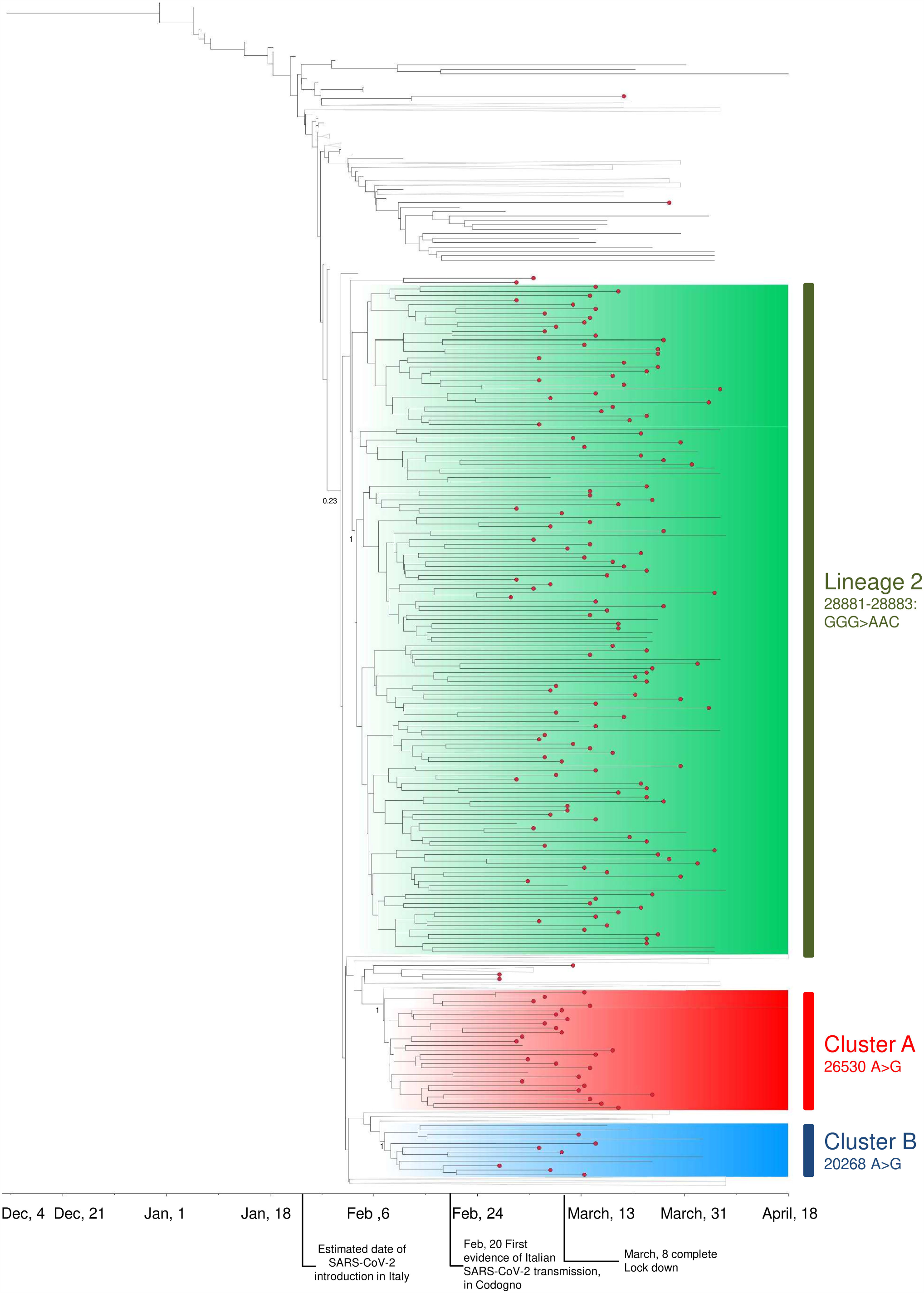
Time-scaled maximum clade credibility tree with SARS-CoV-2 sequences from Lombardy (red circle) and other countries (no circle). SARS-CoV-2 sequences with a posterior probability of 1.00 were highlighted in green (lineage 2), in red (cluster A), in blue (cluster B). All nodes with posterior probabilities <0.8 have been collapsed. Gisaid sequences come from Central Italy (n=6), East Europe (n=6), North Europe (n=6), South America (n=6), Africa (n=7) Japan (n=7), Oceania (n=7), West Asia (n=7), North Italy (n=9), South Asia (n=9), Central Europe (n=11), East Asia (n=11), The Netherlands (n=12), South East Asia (n=13), North America (n=18), British Countries (n=26), China (n=36).

According to the topology of the ML tree, the genetic distance from SARS-CoV-2 reference strain was lower for isolates in Lineage 1 respect to isolates in Lineage 2 (2.7×10^−4^ [2.0×10^−4^;3.7×10^−4^] vs 3.4×10^−4^ [2.7×10^−4^;4.1×10^−4^], P<0.001, Supplementary Figure 3B), thus indicating a closer genetic relatedness with original strains for Lineage 1 respect to Lineage 2. The mean genetic pairwise distance inside Lineages was broadly comparable (3.9×10^−4^ vs 3.5 ×10^−4^, P=0.118), suggesting similar evolutionary characteristics and replication profiles.

These lineages did not contain viral strains isolated in the first months of the outbreak in China (black branches, no dots); this let us hypothesize a transmission chain not directly involving China (i.e., the country where the pandemic originated). Notably, the most closely related viral isolate that clustered outside such Lineages was isolated in Central Europe, in the second half of January. The 61.0% of Lombardy sequences (N=211) were positioned in Lineage 1, interspersed within sequences from West-North Europe, North and South America, and Asia. Of note, most of isolates collected in the South of Lombardy, mainly in Lodi and Cremona, were involved in this lineage (the 93.7% [30/32] and 88.5% [23/26], respectively). In line with this, sequences from Lodi and Cremona with a collection date between February 20 and 22 were the earliest isolates detected in Lineage 1, together with isolates from Central Europe, the Netherlands and South America, at the end of February (February 25 and 29).

The remaining 37.9% of Lombardy sequences (N=131) composed the Lineage 2, together with sequences from European, South American and Asian countries. As Lineage 1 contained most of sequences from the South of Lombardy, Lineage 2 contained the 61.2% of isolates collected in the North, mainly in Lecco (66.7%), Bergamo (61.1%) and Como (58.8%). In line with this, earliest sequences in Lineage 2 included isolates from Bergamo collected between 25 and 29 February, and sequences from Central and North Europe (February 25 and March 2).

This suggests that the initial spread of viral strains belonging to the two Lineages involved mainly the territory in which the first access occurred, with little communication between the two highly affected areas.

The Bayesian molecular clock analysis estimated a mean evolutionary rate of 1.47 x 10^−3^ subs/site/year (95% HPD, 1.34 × 10^−3^-1.62× 10^−3^). The current low genetic diversity of SARS-CoV-2 genomes worldwide implies a very low posterior probabilities (<0.50) for most of the nodes. Concordantly, SARS-CoV-2 sequences in Lineage 1 did not form a monophyletic group (Figure 4). Nevertheless, it was possible to identify 2 transmission clusters with posterior probability support of 1 (Cluster A and B), characterized by the SNP A26530G in M (intra-patient prevalence ∼98%) and A20268G in nsp15 (intra-patient prevalence ∼54%), respectively. In both clusters, Lombardy sequences (24 for Cluster A [Bergamo: 9, Lecco: 5, Milan: 5; Como: 3; Lodi: 1; Cremona: 1], and 6 for Cluster B [Pavia: 2; Como: 1; Milan: 1; Cremona: 1; Brescia: 1]) were intermixed with sequences sampled by other countries (Europe for Cluster A, Europe and West Asia for Cluster B). This further supports the hypothesis that Lineage 1 viruses might be imported into Lombardy on multiple occasions.

Differently by Lineage 1, the Lineage 2 was characterized by a posterior probability of 1 in the maximum credibility tree (i.e., SARS-CoV-2 sequences grouped monophyletically in 100% of trees in the posterior sample, Figure 4). Two SNPs in N (28881-28883:GGG>AAC) characterized the 131 sequences involved in Lineage 2. Of note, these mutations were firstly identified in China, and then spread in all countries, reaching the 37.6% in Italy (intra-patient prevalence:100%) (Figure 2A). Single nucleotide polymorphisms (SNPs), the non-syn A26530G in M (intra-patient prevalence ∼98%) and the syn A20268G in nsp15 (intra-patient prevalence ∼54%), firstly identified in South Asia and Europe (Figure 2A), characterized sequences in Cluster A by Cluster B.

From the molecular clock analysis, we were able to estimate the times of the most recent common ancestor (tMRCA) of all monophyletic traces. Lineage 2 has earlier tMRCAs that coincide with 24^th^ January 2020 (95% HPD 20 January to 28 January 2020). Clusters A and B have few days later tMRCAs, around 27^th^ January 2020 (95% HPD 23 January to 31 January 2020) and 31^th^ January 2020 (95% HPD 26 January to 5 February 2020) (Figure 4). By looking at the distribution of Lombardy sequences in the Bayesian Tree, it is thus conceivable that the circulation of SARS-CoV-2 started in this region with at least two different and nearly contemporaneous foci in the second half of January, one month before the first COVID-19 diagnosis in Codogno (Lodi). This is in line with evidences of initial outbreaks in other European countries, including Germany, in the second half of January.^13^

The Bayesian reconstruction also allowed to estimate the median tMRCA estimate of the COVID-19 pandemic that was 4 December 2019 (95% HPD 29th November to 21th December 2019; Figure 4), consistent with previous estimates.^14-15^

## Discussion

These data on the genomic epidemiology of SARS-CoV-2 in Lombardy, based on the largest number of whole genome SARS-CoV-2 sequences generated in a single study, indicate the simultaneous circulation of two lineages of SARS-CoV-2 likely starting from multiple introductions, which occurred in the second half of January (Figure 3 and 4), one month before the first COVID-19 case detected in Codogno (Lodi). Of note, the ML tree showed that these two chains of transmission were wide in size and fast in spreading.

The MRCAs of these two SARS-CoV-2 lineages were closely related in time, as they dated just one month before the first diagnosed case, yet they showed different preferential geographical circulations. Lineage 1 mostly interested the Southern Lombardy, including Lodi and Cremona (Table 1), while Lineage 2 predominated in the North of Lombardy, mostly in Bergamo and its adjacent territories (such as Alzano and Nembro, that represented a major focus of the epidemics). The predominance of these lineages in different territories, the lack of a monophyletic signal for Lineage 1, and the detection of two well supported clusters inside it (Figure 4), supports the hypothesis of multiple and dislocated introductions of SARS-CoV-2 with different route of viral transmission. These data are in line with recent description of dynamics of the SARS-CoV-2 pandemic in densely populated areas,^14,16^ where SARS-CoV-2 started its spread by multiple, independent, and frequently undetected, introductions.

Altogether, the spread of two lineages in the two most populated cities of the Lombardy region (Milan and Pavia), along with a broadly comparable level of genetic pairwise distance, and similar viral load levels within them, support their similar fitness, and hence exclude the occurrence of a strong competition between them. Consistent with this, using a Chi-squared test, no trend of association between disease severity (including evidence of interstitial pneumonia, and severe presentation) and lineages was detected (Table 1). Nevertheless, if specific clades or mutational variants might modulate the clinical presentation and spread of the disease should be prudently evaluated, until new and extensive data focused on this setting will be available.

At this regard, sequence analysis defined the presence of founding mutations characterizing these two transmission chains. Lineage 2 is defined by sequences characterized by the 3 SNP 28881-28883:GGG>AAC, resulting in two amino acid 203-204:RG>KR changes in the nucleocapsid (N) protein of the SARS-CoV-2. These mutations are localized at the end of serine-arginine (SR) dipeptide of the SR-rich motif (aa 183-195: SSRSSSRSRNSSR [NSTPGSS<u>RG]</u>) characterizing the N protein of SARS-CoV-2, and introduce a lysine between a SR-dipeptide. By previous study on SARS-CoV, which shares the 75% amino acid similarity with SARS-CoV-2, the deletion of the SR-rich motif, or its mutation, significantly reduce viral genomic transcription, the levels of the infectious virions, and the rate of host cell translational inhibitory activity.^17,18^ Whether the 203-204:RG>KR changes would be able to affect SARS-CoV and/or SARS-CoV-2 virus replication is intriguing, yet currently unknown.

Cluster A and B, in Lineage 1, are characterized by the presence of two SNPs, the non-syn A26530G (intra-patient prevalence ∼98%), leading to the D3G mutation within a B-cell epitope of M protein^19^, and the syn A20268G in nsp15 (intra-patient prevalence ∼54%).

Looking at the overall variability of SARS-CoV-2, we found that only 7 SNPs (2 out 7 synonymous) characterized our consensus sequences, highlighting a good conservation rate of this virus along time. This conservation rate is confirmed within the spike structural protein, where only 2 mutations, one of them at low prevalence (i.e, C23575T, corresponding to the amino acid variant T671I), were detected. None of these mutations have a role in altering pre-existing N-glycosylation sites or in creating new ones,^20^ which may be beneficial for the development of vaccines strategies.

Worth of mention is the SNP A to G at position 23403, corresponding to the variant D614G in spike protein, detected in the 67.8% of our SARS-CoV-2 sequences. As already known, this variant is observed frequently in European countries, such as the Netherlands, Switzerland, and France, but seldom observed in China. Its rapid fixation at population level might suggest a role in viral entry, and enhancement of interaction between receptor-binding-domain of the S protein with the entry receptor ACE2. This variant, located within a B-epitope, causes the substitution of a large acidic residue (aspartic acid), with a small hydrophobic residue (glycine). Further investigations are required to define if such marked differences in both size and hydrophobicity in the middle of the epitope might compromise the binding affinity to antibodies against wild-type spike protein, elicited by vaccines.^21^

Our study may have some limitations. The analysis of phylogenetic structures during such an early phase of the pandemic should be interpreted carefully, as the number of mutations that define phylogenetic lineages is small and may be similar to the rate of potential errors introduced during reverse transcription, PCR amplification, or sequencing.^22^ To overcome these problems, Bayesian approach, known to be a powerful way to estimate species divergence, and thus expected to provide more robust results, was applied. Moreover, the integration of host characteristics (such as geographical location, collection date and clinical manifestations) aided phylogenetic interpretation. Moreover, the intra-host variability of SARS-CoV-2, and the role of potential existing minority variants has not been investigated here. Initial evidences suggest that intra-host variation of SARS-CoV-2 can be frequently found among clinical samples (median number of intra-host variants: 1–4), but at the same time these variants were not observed in the population as polymorphisms, probably suggesting a bottleneck or purifying selection involved.^23,24^ Thus, ad hoc designed studies are necessary to provide an extensive overview of SARS-CoV-2 intra-host variability and minority variants description, if and how these minority variants can spread in the population, and their potential role in virulence and transmissibility.

In the peak of epidemic, SARS-CoV-2 diagnosis was mainly addressed to symptomatic cases or subjects at high risk of exposure (i.e. health care workers exposed to positive patients without adequate protection). This approach may have caused a substantial underestimation of positive subjects, preventing to include in our analysis asymptomatic infections, whose role in the transmission chains and in influencing the evolution of epidemic remains a challenge to investigate. Moreover, even if the number of samples here analysed is noteworthy, the epidemic in the East (i.e. Brescia and Mantua) and the valleys of the North (i.e. Valtellina, and Valcamonica, Figure 1) is poorly represented. With exception of Brescia, that represents today one of the most COVID-19 affected area after Bergamo, Cremona and Lodi, Mantua and the valleys account today for the 6.7% of confirmed COVID-19 cases in Lombardy, a percentage that does not represent a major issue for the good reproducibility of SARS-CoV-2 epidemic in the region.

In conclusion, this study allows the identification of SARS-CoV-2 lineages circulating in the most affected COVID-19 area at the beginning of 2020, representing a huge reserve of genetic information of a virus that became able to pandemic spread, and caused in Lombardy more than 16,000 deaths in some weeks. We cannot exclude that this multiple and simultaneous circulation of SARS-CoV-2 strains can have exacerbated the transmissibility potential of the virus and thus create a real viral storm in such high densely populated region. Only the large-scale surveillance and intervention measures implemented in the early March in Italy were effective in reducing community transmission, ultimately containing the epidemic and limiting the dissemination to other regions.

## Data Availability

Corresponding author is Carlo Federico Perno, MD, PhD, University of Milan. The data that support the findings of this study are available from the corresponding author upon reasonable request.

## Acknowledgments

We thank Biodiversa s.r.l. for providing technical support, particularly Dr. Marco Dotto, for his valuable assistance in the implementation of the study. The authors also thank Dr. Silvia Nerini and all the staff of the Microbiology and Virology Laboratory of ASST Grande Ospedale Metropolitano Niguarda and IRCCS San Matteo for outstanding technical support in processing swab samples, performing laboratory analyses and data management.

## Author contributions

CA, VCe, AP equally contributed in study design, data collection, data analysis, data interpretation, writing; VCo performed bioinformatic analysis and data processing; MT, LC, SC, SR, FG, FN processed samples and collected data; SG helped in bioinformatic analysis; EM and MA helped in sample processing and data collection; CV, RF, OME, MP recruited samples and enrolled patients; CFP and FB conceived and directed the study, and critically revised the manuscript.

